# Genetically predicted plasma erythropoietin levels and clonal haematopoiesis risk: a Mendelian randomisation study

**DOI:** 10.1101/2024.11.19.24317548

**Authors:** George Richenberg, Paul Carter, Tom R. Gaunt, Siddhartha P. Kar

**Affiliations:** MRC Integrative Epidemiology Unit, University of Bristol, Bristol, UK; Population Health Sciences, Bristol Medical School, University of Bristol, Bristol, UK; Victor Phillip Dahdaleh Heart and Lung Research Institute, University of Cambridge, Cambridge, UK; Early Cancer Institute, University of Cambridge, Cambridge, UK

## Abstract

The aetiology of clonal haematopoiesis (CH) and its progression to haematological malignancies is poorly understood. Erythropoietin (EPO) has been linked to favourable haematopoietic stem and progenitor cell clonal compositions in mice and humans, but a direct large-scale assessment of its role in CH is lacking. We evaluated the association between plasma EPO levels and overall, *DNMT3A*- and *TET2*-mutant CH risks using Mendelian randomisation (MR). Germline variants in the EPO gene (common promoter variant rs1617640 and rare missense mutation rs11976235) from a genome-wide association study (GWAS) of plasma EPO levels in 33,657 individuals were used to genetically predict EPO levels. CH associations were obtained from a European ancestry GWAS of 25,657 CH cases, including 16,219 *DNMT3A*- and 3,918 *TET2*-mutant CH cases, and 342,869 “controls” without detectable CH. MR analyses using the Wald ratio revealed that higher plasma EPO levels proxied by rs1617640 were associated with reduced risks of overall (odds ratio/OR (95% confidence interval/CI)=0.68 (0.50-0.93); P=0.01), *DNMT3A*- (OR (95% CI)=0.68 (0.48-0.96); P=0.03) and *TET2*-mutant (OR (95% CI)=0.48 (0.24-0.97); P=0.04) CH. These point estimates were directionally consistent with those obtained using the rare variant rs11976235, and in analyses of overall CH in African, East Asian and South Asian ancestry individuals. Bayesian evaluation also indicated that elevated plasma EPO levels were protective for *DNMT3A*- and *TET2*-mutant CH with probabilities ≥92% and 73%, respectively. Thus, increased genetically predicted plasma EPO levels are protective for overall CH and its two most common subtypes. Our findings suggest that pharmacologically raising EPO may be a promising strategy worth further investigating for the prevention of CH and possibly to prevent its progression to haematological malignancies in high-risk individuals.

## INTRODUCTION

Clonal haematopoiesis (CH) is an age-related condition characterised by the accrual of somatic driver mutations in haematopoietic stem and progenitor cells (HSPCs) that confer them with a selective advantage leading to their clonal expansion^1-3^. The prevalence of CH increases with age and is detectable in 10 to 20% of individuals over the age of 70 years. CH is associated with an increased risk of haematological malignancies, cardiovascular disease and mortality^1^. Somatic loss-of-function mutations in the *DNMT3A* and *TET2* genes, respective regulators of DNA methylation and de-methylation, are the most prevalent drivers of CH accounting for nearly two-thirds of CH cases^4, 5^. Although cancer treatment, inflammation and smoking, have been identified as risk factors for CH, there remain substantial gaps in our understanding of the aetiology of CH^6^.

Erythropoietin (EPO) is a glycoprotein hormone produced by the kidneys that stimulates erythropoiesis in response to hypoxia to promote red blood cell production, and consequently increase blood oxygen levels^7, 8^. Beyond this well-defined role, EPO has been shown to influence the relative composition of HSPC clones in murine models facilitating the expansion not just of erythroid cells but also having a direct effect on the myeloid compartment through multipotent progenitor populations^9^. Recent research in humans has also shown that HSPCs derived from cord blood and adult healthy bone marrow and genetically engineered *in vitro* to carry known leukaemogenic *DNMT3A* mutations did not expand in response to EPO, while EPO stimulation exerted a selective effect spurring expansion of HSPCs with non-pathogenic *DNMT3A* mutations^10^. Despite these emerging links between EPO and HSPCs, the association between EPO levels and CH susceptibility is yet to be directly evaluated. This unexplored area presents a potential opportunity to offer fresh insight into the aetiology and potential prevention of CH.

In the absence of randomised control trials (RCTs), Mendelian randomisation (MR) serves as a powerful epidemiological study design to assess whether an exposure has a causal effect on an outcome of interest^11^. MR uses germline genetic variants as instrumental variables (IVs) to proxy the exposure of interest, akin to the random assignment process in RCTs^11^. The random segregation and independent assortment of alleles at conception and the fact they remain fixed thereafter minimises the susceptibility of MR to confounding and reverse causation overcoming the major limitations of observational epidemiology^11^. While recent MR studies have offered valuable insights into CH risk factors^4, 12^, there is substantial additional potential to use MR to identify novel and modifiable exposures associated with CH risk.

In this study, we used MR to test for associations between genetically predicted plasma EPO levels (exposure) and CH risk (outcome). Germline variants associated with plasma EPO levels were identified from a genome-wide association study (GWAS) of plasma EPO levels in over 33,000 individuals of European ancestry in the UK Biobank^13^. We used two variants in the *EPO* gene (i.e., *cis*-protein quantitative trait loci (pQTLs)) that were identified in this GWAS as separate IVs to proxy EPO levels. Germline variants located in or near the gene coding for a protein (cis-pQTLs) are more likely to directly impact the protein’s transcription or translation, making them more biologically relevant for instrumenting specific targets such as EPO^14, 15^. These included an *EPO* promoter variant associated with plasma EPO levels that has been functionally validated via genome editing as a regulator of EPO gene expression and has been previously leveraged in “drug target” MR studies of EPO^16^, and a rare missense variant in *EPO* that was identified as the index variant in the *EPO* region in a recent of GWAS of plasma EPO levels^13^. Summary statistics for the association of these with CH were obtained from the largest published GWAS of CH susceptibility comprising over 368,000 individuals of European ancestry and from GWAS of CH in individuals of African, East Asian, and South Asian ancestry – all from the UK Biobank^5^. We adopted MR with a two-sample design to examine associations between genetically predicted EPO levels and risks of overall, *DNMT3A*-mutant and *TET2*-mutant CH (Figure 1). We also re-analysed the MR results using a Bayesian approach to interpret our findings within a probabilistic framework and performed a number of additional analyses to evaluate their robustness (Figure 1).

**Figure 1.**
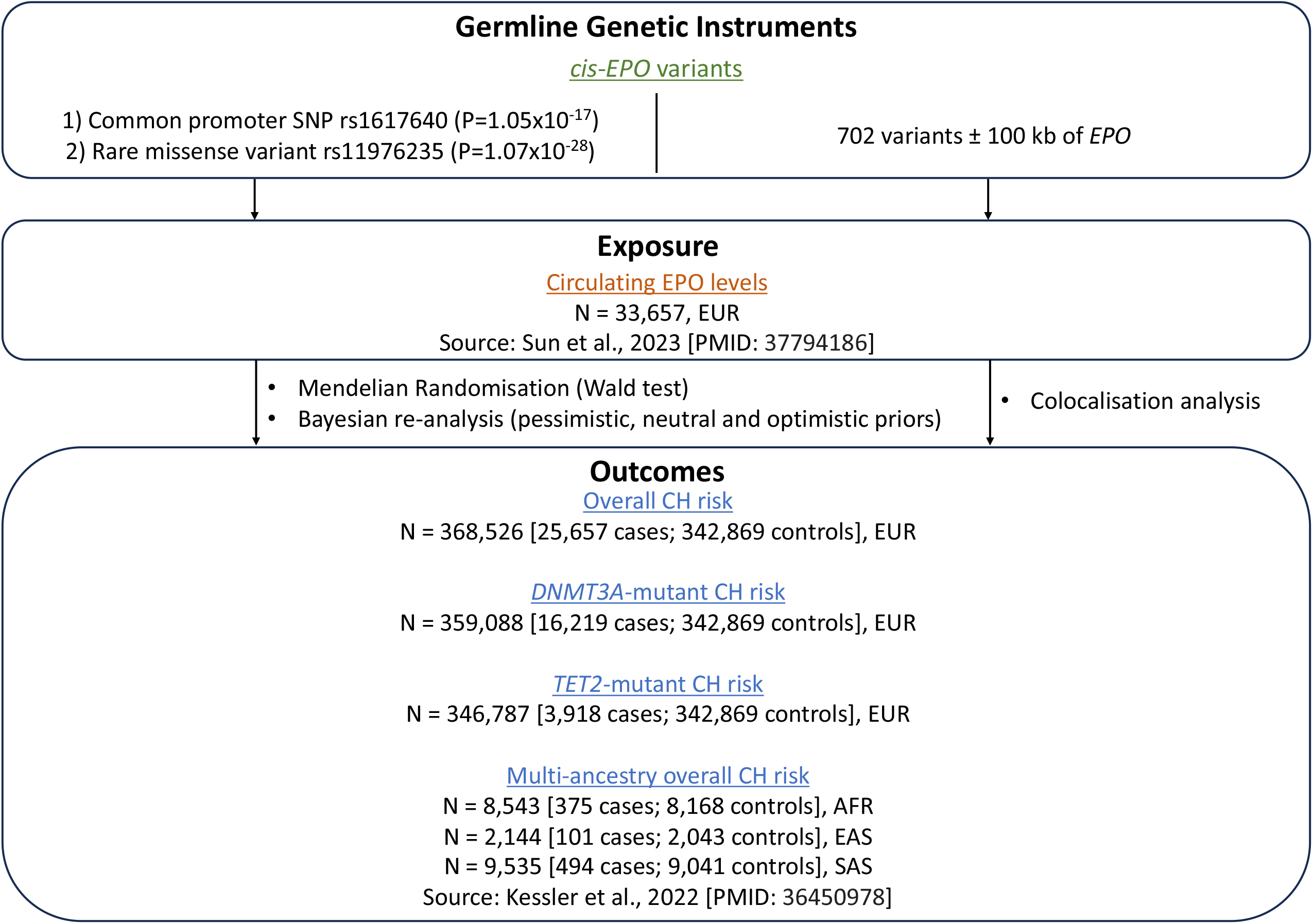
Schematic overview of the study. Abbreviations: EUR = European, AFR = African, EAS = East Asian, and SAS = South Asian ancestries.

## METHODS

### Study design

We used two-sample Mendelian randomisation to examine associations between genetically predicted plasma erythropoietin levels and clonal haematopoiesis risk using summary genetic association data from published GWAS of circulating EPO and CH. MR uses germline variants associated with an exposure as instruments or instrumental variables to proxy that exposure and assess for the causality of associations between the exposure and outcome^11^. Germline variants can be defined as instruments for an exposure if they satisfy three assumptions: (i) the relevance assumption (i.e., the variants are associated with the exposure of interest), (ii) the exchangeability assumption (i.e., the association between the variants and the outcome is independent of confounding) and (iii) the exclusion-restriction assumption (i.e., the variants influence the outcome solely through their effects on the exposure)^17^. We also undertook a Bayesian re-analysis of our MR findings to provide a probabilistic interpretation of the findings. Finally, we rigorously evaluated the impact of genetic confounding, pleiotropy, and sample overlap on our results.

### Data sources

#### Erythropoietin data

Summary statistics for germline variants associated with plasma erythropoietin levels were obtained from a GWAS of 33,657 individuals of European ancestry in the UK Biobank^13^. We used the common single nucleotide polymorphism (SNP) in the *EPO* promoter, rs1617640 (minor allele frequency (MAF) = 0.40), as our primary instrumental variable and the rare missense *EPO* variant, rs11976235 (MAF = 0.006), as a distinct secondary instrumental variable to proxy plasma EPO levels for MR analysis. Both variants were independent in terms of linkage disequilibrium (*r*^2^ = 7 × 10^−4^). The SNP rs1617640 was first identified as being associated with plasma EPO protein levels in a GWAS meta-analysis of 6,127 individuals of European and African American ancestry by Harlow, *et al*.^16^. In the same study, which was not based on the UK Biobank, the authors used a knock-in cell model generated by CRISPR-Cas9 single-base gene editing to demonstrate allele-specific regulation of *EPO* gene expression levels by rs1617640. In addition to this functional evidence, the same SNP was subsequently found to have a genome-wide significant (P = 1.05 × 10^−17^) association with plasma EPO levels in the UK Biobank GWAS^13^. The rare missense EPO gene variant, rs11976235, was the strongest association for EPO protein levels in the EPO gene region in the UK Biobank (P = 1.07 × 10^−28^).

### Clonal haematopoiesis data

Summary statistics for germline genetic variants associated with CH risk were obtained from a GWAS of 368,526 individuals of European ancestry in the UK Biobank^5^. This GWAS included 25,657 individuals with CH overall (including 16,219 with *DNMT3A*-mutant and 3,918 with *TET2*-mutant CH), and 342,869 individuals without any detectable CH. CH phenotyping was based on blood (buffy coat) DNA exome sequencing data focused on 23 specific genes and is described in detail in Kessler, *et al*^5^. We also used the UK Biobank-based GWAS summary statistics for overall CH risk in individuals of African (375 cases/8,168 controls), East Asian (101 cases/2,043 controls) and South Asian (494 cases/9,041 controls) ancestries available from Kessler, et al^5^.

### Statistical analyses

#### Mendelian randomisation

We performed two-sample MR analyses using the TwoSampleMR R package (version 0.5.7)^18^ to test for associations between genetically predicted plasma EPO levels and overall, *DNMT3A*- and *TET2*-mutant CH risks. Summary genetic association data from the exposure and outcome traits were matched by variant chromosome and position and the direction of effect size estimates (beta coefficients) harmonised to refer to the same effect allele. The MR association between exposure and outcome was calculated using the Wald ratio with standard errors for the Wald ratio approximated by the delta method^19^. Instrumental variable strength was assessed using the F-statistic to assess the relevance assumption of MR, with F < 10 considered as evidence of weak instruments^20^.

### Bayesian re-analysis of Mendelian randomisation findings

We also re-evaluated our MR results in a Bayesian context to yield intuitive and easily interpretable probabilities that EPO had a protective effect on CH risk. To do this, we used the standardised framework described in detail in Zampieri, *et al*^21^. Briefly, we modelled the MR results under three priors: strongly neutral, moderately pessimistic, and weakly optimistic (to use the terms and prior probabilities (see below) used by Zampieri, *et al*^21^) reflecting *a priori* neutrality/pessimism/optimism towards the fact that EPO had a protective effect on CH. All three priors were normally distributed. The strongly neutral prior was centred on an odds ratio of 1 with a standard deviation (SD) of 0.205, to reflect a prior probability of a protective effect of 50%. The moderately pessimistic prior was centred on an odds ratio of 1.5 with a SD of 0.3247, to reflect a prior probability of a protective effect of 15%. The weakly optimistic prior was centred on an odds ratio of 0.8 with a SD of 0.4255, to reflect a prior probability of a protective effect of 70%. Posterior median probabilities and corresponding 95% credible intervals were calculated based on these priors, incorporating the MR results as the likelihoods. We further presented the posterior probabilities modelled under each of the three priors in two ways: (i) posterior probability that the MR odds ratio (OR) for the effect of EPO on CH was < 1 (indicating at least some protective effect) and (ii) posterior probability that the MR OR was < 0.8 (indicating a protective effect > 20%).

### Colocalisation analysis

Colocalisation analysis was used to assess whether plasma EPO levels and CH risk associations at the *EPO* gene region (i.e., ±100 kb of *EPO*) share the same underlying genetic association signal or are underpinned by distinct signals that confound the MR result due to the effect of linkage disequilibrium. Evidence for colocalisation assesses the exchangeability assumption of MR. We performed colocalisation using the coloc R package (version 5.2.2) with the default recommended priors^22^. Colocalisation calculates approximate Bayes factors to compute five posterior probabilities: neither trait has a genetic association in the region (PP.H0), only EPO has a genetic association in the region (PP.H1), only CH has a genetic association in the region (PP.H2), both traits have distinct genetic associations in the region (PP.H3), and both traits share the same genetic association in the region (PP.H4)^22^.

If both PP.H3 and PP.H4 are low, it suggests that the power of colocalisation analysis is low. In MR analyses, the genetic variant selected to proxy an exposure is usually genome-wide significant (P < 5 × 10^−8^). Further, we only used colocalisation analysis to follow up MR findings that achieved P < 0.05. In such a situation if PP.H1 is high and close to 1 while PP.H2 is low and close to 0, it indicates that the outcome data set is underpowered for colocalisation. Given such a situation the alternative colocalisation probability (altPP.H4) of PP.H4/(PP.H3 + PP.H4) that provides the posterior probability of colocalisation conditional on the presence of an association with the outcome provides a more powerful measure of the relative probability of a shared versus distinct genetic association for exposure and outcome in the region^23^. Moreover, application of the alternative colocalisation probability is consistent with the use of colocalisation analysis in the MR setting to rule out confounding by distinct genetic associations in the region. Therefore, we calculated altPP.H4 for all exposure-outcome pairs that yielded MR P-values < 0.05.

### Additional analyses

We searched for rs1617640 and rs11976235, the two variants used to proxy plasma EPO levels, in the PhenoScanner database^24^ to identify additional associations at P < 5 × 10^−8^ between these variants and other diseases and traits. This was done to identify potential pleiotropic associations for these SNPs that could confound the MR estimate between EPO and CH.

The EPO GWAS that we used for our MR analyses was based on a subset of the UK Biobank. The CH GWAS that we used was also based on the UK Biobank. To rule out the possibility that the MR associations we identified were being driven by bias from this sample overlap^25^, we also re-calculated the MR results for rs1617640 using variant-EPO genetic association summary statistics from the GWAS meta-analysis of plasma EPO levels reported by Harlow, *et al*^16^. As described above (under “Erythropoietin data”) the study by Harlow, *et al* was based on 6,127 individuals who were not part of the UK Biobank^16^.

All analyses were conducted in R (version 4.3.0) and have been reported in accordance with the “strengthening the reporting of observational studies in epidemiology using Mendelian randomisation” (STROBE-MR) guidelines^26^. All plots were generated using the ggplot2 R package (version 3.4.3).

## RESULTS

This two-sample Mendelian randomisation study examined the association between *cis*-genetically predicted plasma erythropoietin levels and clonal haematopoiesis risk (Figure 1). Outcome data on CH were obtained from a GWAS of 25,657 individuals with CH, including 16,219 with *DNMT3A*-mutant and 3,918 with *TET2*-mutant CH, and 342,869 individuals without detectable CH^5^. A genome-wide significant (P < 5 × 10^−8^), common (MAF = 0.40), variant in the EPO promoter, rs1617640, obtained from a GWAS of plasma EPO levels in 33,657 individuals^13^ was used as the primary instrumental variable to proxy EPO as an exposure for the MR analyses. We also separately evaluated a second instrumental variable comprising a rare (MAF = 0.006) missense variant, rs11976235, from the same GWAS^13^. F-statistics for rs1617640 (F = 73) and rs11976235 (F = 124) were > 10 suggesting a low likelihood of weak instrument bias and satisfying the relevance assumption of Mendelian randomisation.

### Two-sample Mendelian randomisation

The A allele of rs1617640 (beta (95% CI) = 0.07 (0.05 – 0.08); P = 1.05 × 10^−17^) and the T allele of rs11976235 (beta (95% CI) = 0.55 (0.45 – 0.64); P = 1.07 × 10^−28^) were associated with increased plasma EPO levels (Figure 2A). We found that an increase in genetically predicted plasma EPO level, proxied by the common variant rs1617640, was associated with reduced risks of overall CH (OR (95%CI) = 0.68 (0.50 – 0.93); P = 0.01), *DNMT3A*-mutant CH (OR (95%CI) = 0.68 (0.48 – 0.96); P = 0.03) and *TET2*-mutant CH (OR (95%CI) = 0.48 (0.24 – 0.97); P = 0.04) (Figure 2B). These results were consistent in terms of the direction of the point estimate of the effect size and had overlapping confidence intervals with those obtained from proxying an increase in EPO using the rare variant rs11976235: overall CH (OR (95% CI)= 0.87 (0.69 – 1.11); P = 0.26), *DNMT3A*-mutant CH (OR (95% CI) = 0.81 (0.62 – 1.06); P =0.12) and *TET2*-mutant CH (OR (95% CI) = 0.79 (0.46 – 1.36); P = 0.40) (Figure 2B). The MR results were also consistent in terms of the direction of the point estimate of the effect size (but with much wider confidence intervals due to the substantially smaller sample sizes) with those obtained using rs1617640 in individuals of African (OR (95% CI) = 0.32 (0.02 – 4.22); P = 0.39), East Asian (OR (95% CI) = 0.39 (0.00 – 188.15); P = 0.77), and South Asian (OR (95% CI) = 0.60 (0.07 – 5.32); P = 0.64) ancestries (Figure 3).

**Figure 2.**
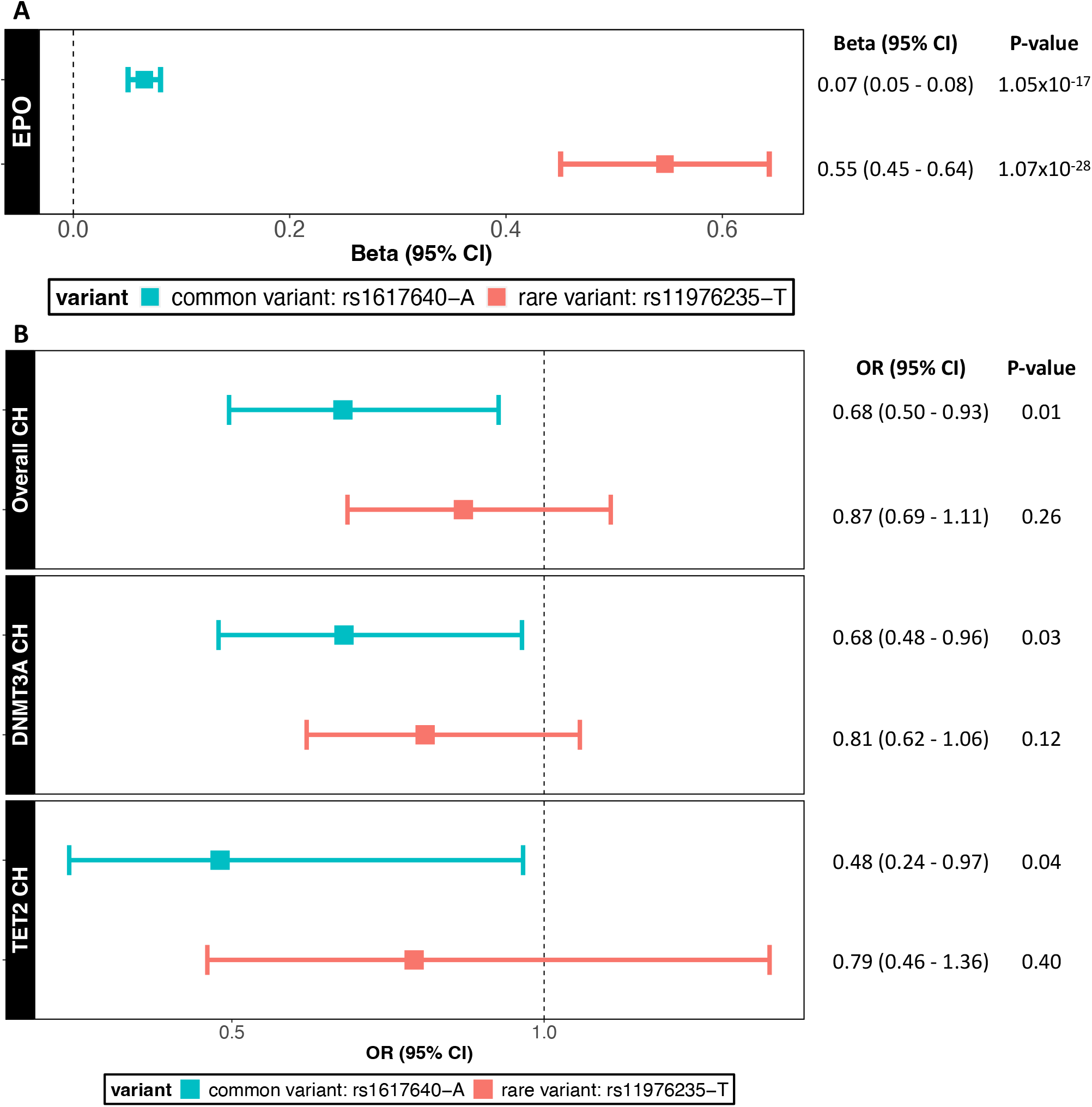
Associations between the variants rs1617640 and rs11976235 and plasma EPO levels (A) and MR results for the association between EPO and overall, *DNMT3A*-, and *TET2*-mutant CH risks (B). OR = odds ratio; CI = confidence interval.

**Figure 3.**
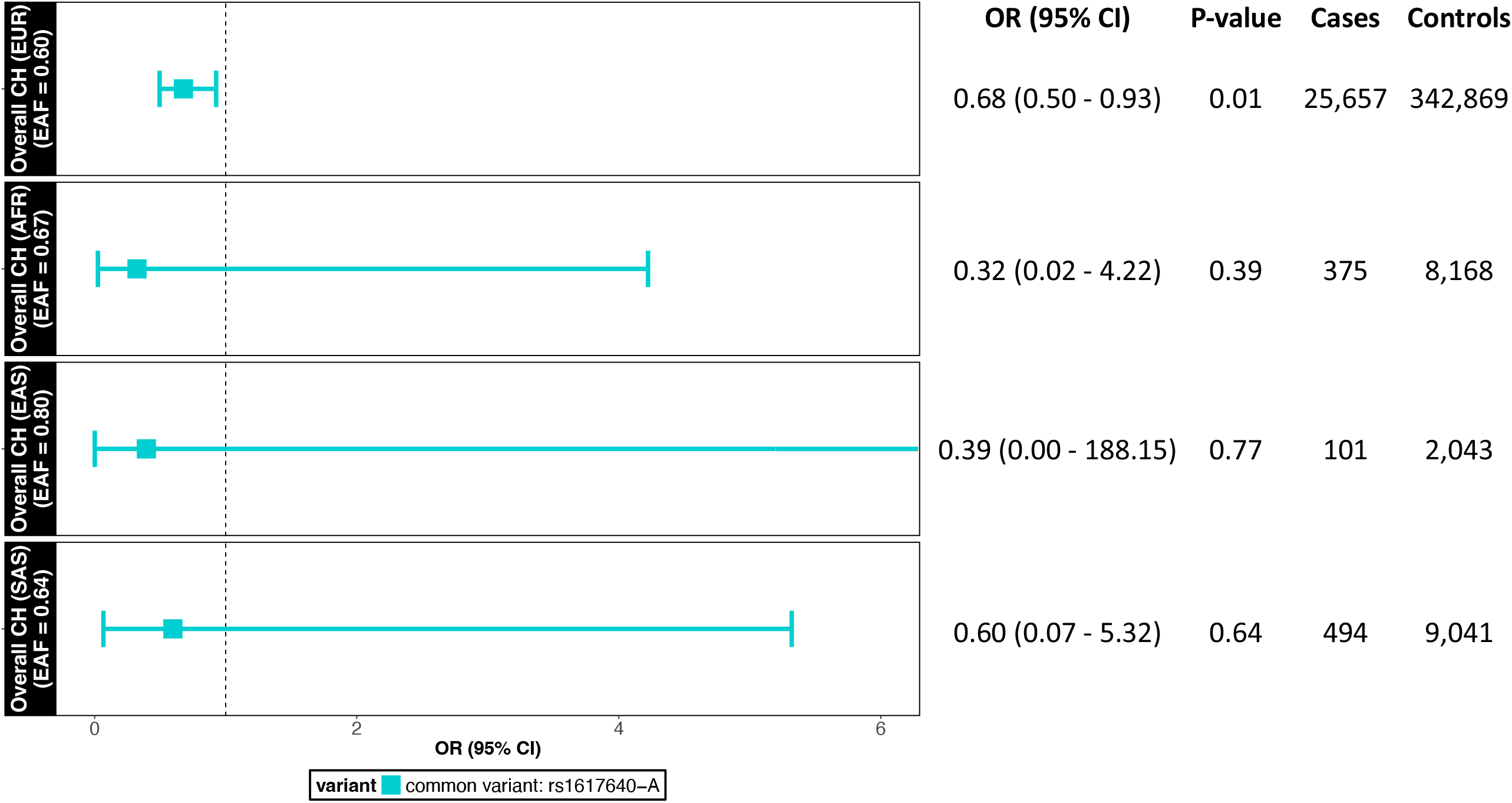
MR results for the association between plasma EPO levels as proxied by rs1617640 and overall CH risk in individuals of European (EUR), African (AFR), East Asian (EAS), and South Asian (SAS) ancestries.

### Bayesian re-analysis of Mendelian randomisation findings

We performed a Bayesian re-analysis of our *DNMT3A*- and *TET2*-mutant CH MR results using the standardised framework described by Zampieri, *et al*^21^. This was done to provide a probabilistic interpretation of these results in the context of pessimistic, neutral/sceptical, and optimistic prior beliefs about the association between EPO and CH, rather than relying solely on a rigid, binary interpretation of P-values at the P = 0.05 threshold to declare evidence of an association.

We used the three priors to calculate the posterior probabilities that genetically predicted elevated plasma EPO levels were protective for *DNMT3A*-mutant CH and *TET2*-mutant CH to some extent (OR < 1) or with a protective effect size of at least 20% (OR < 0.8). Under all priors, we found high posterior probabilities (92.4% - 98.7%) that increased EPO reduced *DNMT3A*-mutant CH risk (OR < 1) and modest probabilities (49.6% - 80.3%) that increased EPO reduced *DNMT3A*-mutant CH risk by at least 20% (OR < 0.8) (Figure 4A-C). There were also high posterior probabilities (72.7% - 97.1%) that increased EPO reduced *TET2*-mutant CH risk (OR < 1) and low-to-modest probabilities (37.3% - 86.2%) that increased EPO reduced *TET2*-mutant CH risk by at least 20% (OR < 0.8) across all priors (Figure 5A-C).

**Figure 4.**
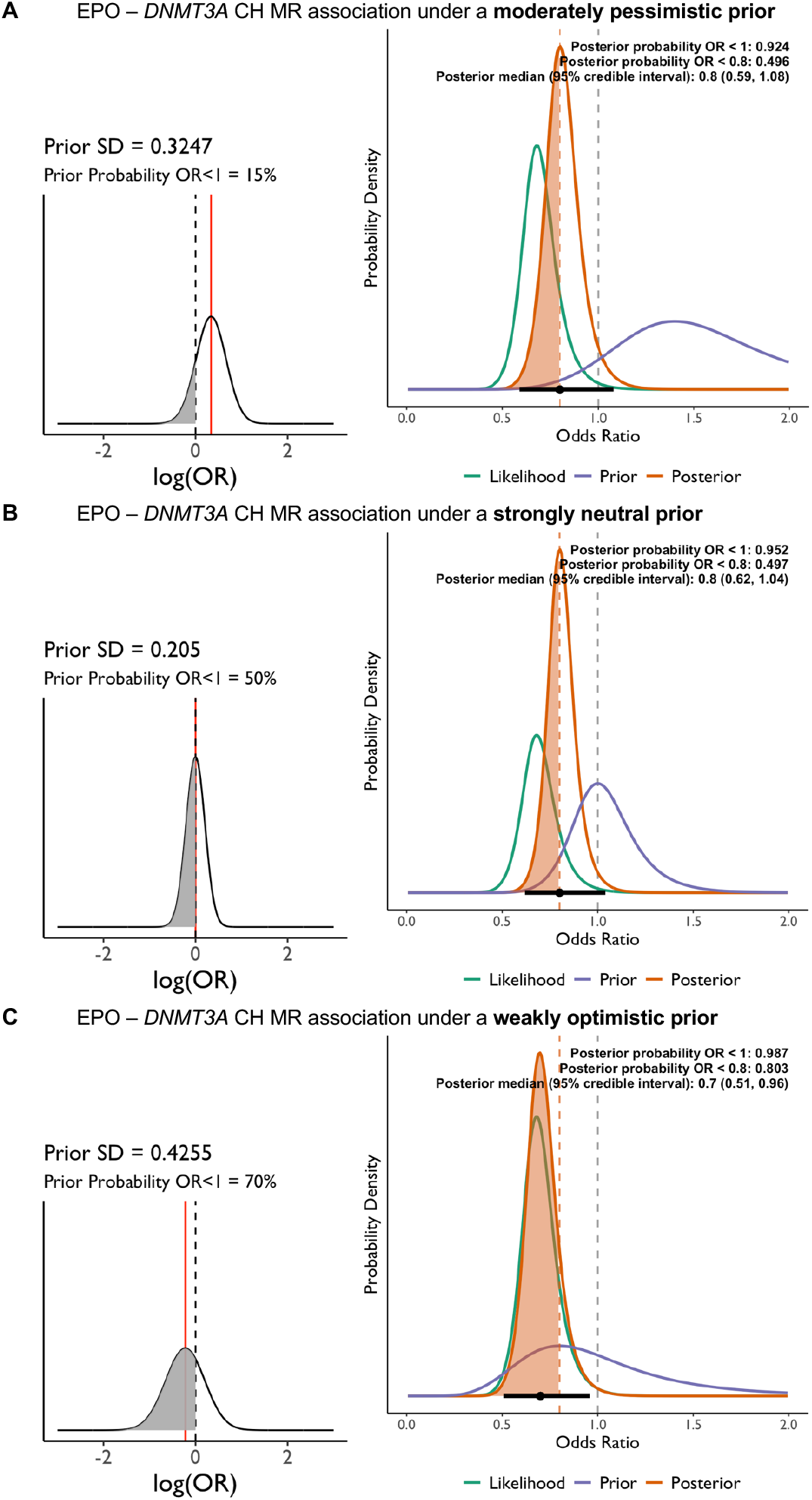
Bayesian re-analyses of MR results for the EPO-DNMT3A CH association under pessimistic (A), neutral (B), and optimistic (C) priors. OR = odds ratio; SD = standard deviation.

**Figure 5.**
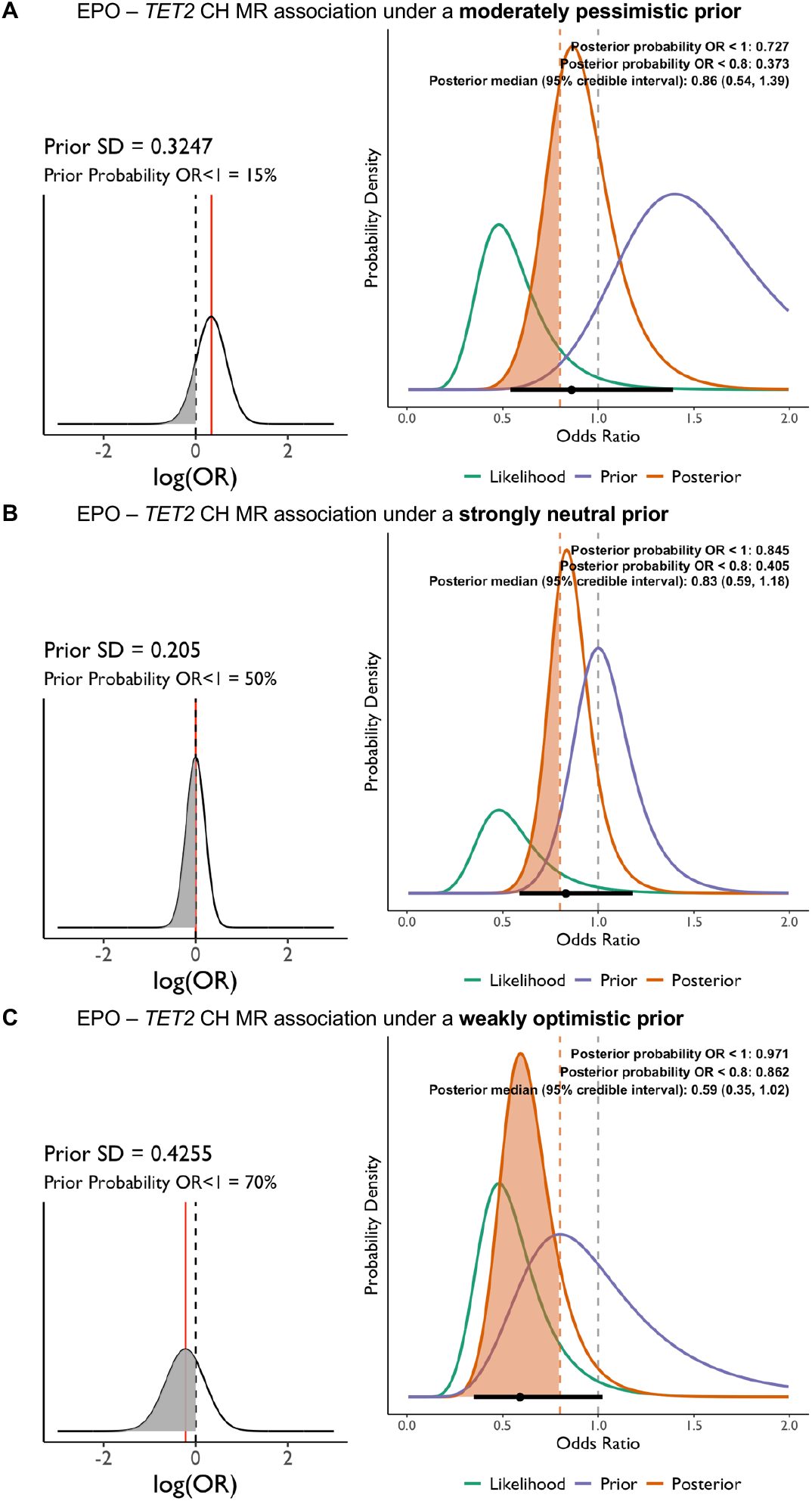
Bayesian re-analyses of MR results for the EPO-TET2 CH association under pessimistic (A), neutral (B), and optimistic priors. OR = odds ratio; SD = standard deviation.

### Colocalisation analyses

We performed colocalisation analyses to explore the possibility of genetic confounding of the Mendelian randomisation results from the effects of linkage disequilibrium. That is, we evaluated the relative probability that either shared or distinct genetic association signals for plasma EPO levels and CH risk at the *EPO* locus were driving the Mendelian randomisation association between EPO and CH. We found that the posterior probabilities indicative of a shared underlying genetic association signal between EPO and CH at the *EPO* locus were higher than the posterior probabilities indicative of distinct signals: overall CH (PP.H3 = 0.04; PP.H4 = 0.05; altPP.H4 = 0.55), *DNMT3A*-mutant CH (PP.H3 = 0.03; PP.H4 = 0.09; altPP.H4 = 0.73), and *TET2*-mutant CH (PP.H3 = 0.04; PP.H4 = 0.07; altPP.H4 = 0.62) (Supplementary Table 1).

### Additional analyses

We used PhenoScanner^24^ to evaluate the association between rs1617640 or rs11976235 and other diseases and traits beyond EPO and CH. As expected for an EPO-associated variant, we found associations with red blood cell count, haematocrit, haemoglobin concentration, mean corpuscular haemoglobin, mean corpuscular haemoglobin concentration, and mean corpuscular volume (Supplementary Table 2). Additional associations identified for rs1617640 were with platelet count and nephropathy as a complication of type 1 diabetes and between rs11976235 and death from appendix-related causes (Supplementary Tables 2 and 3). None of these additional associations were likely to explain the EPO to CH association.

Both exposure (EPO) and outcome (CH) GWAS in our two-sample MR study were based on the UK Biobank. To rule out the possibility of winner’s curse arising from this sample overlap driving the MR estimates, we re-did the analyses after obtaining the variant-EPO association estimates for rs1617640-A from a GWAS of 6,127 individuals that did not include the UK Biobank. Point and interval estimates for the associations between EPO and overall, *DNMT3A*- and *TET2*-mutant CH remained almost identical to those identified in the original analyses (Supplementary Figure 1).

## DISCUSSION

In this Mendelian randomisation study, we investigated the association between germline genetically predicted plasma erythropoietin levels and the risk of clonal haematopoiesis, focusing on overall CH and its two most common subtypes driven by *DNMT3A* and *TET2* mutations. Our findings suggest that higher genetically predicted plasma EPO levels are associated with a reduced risk of CH. Specifically, the common promoter variant rs1617640, which proxies increased plasma EPO levels, was associated with a decreased risk of overall CH, as well as *DNMT3A*-mutant and *TET2*-mutant CH. These associations were consistent in direction and magnitude when using a rare missense variant rs11976235 as an alternative instrument and across different ancestral populations, albeit with wider confidence intervals due to smaller sample sizes.

Our results align with emerging experimental evidence highlighting the role of EPO in haematopoietic stem and progenitor cell dynamics. In murine models, EPO has been shown to influence the clonal composition of HSPCs, promoting not only erythroid but also myeloid progenitor expansion through effects on multipotent progenitor cells^9^. In human studies, EPO stimulation selectively expanded HSPCs harbouring non-pathogenic *DNMT3A* mutations but not those with leukaemogenic mutations, suggesting a differential impact based on the mutational status of HSPCs^10^. The protective association between higher EPO levels and reduced CH risk observed in our study may be attributed to several biological mechanisms. While the evolutionary trajectories and germline genetic susceptibility profiles of *DNMT3A*- and *TET2*-mutant CH are now known to diverge^4, 27^, our results suggest that EPO is protective against both these distinct forms of CH, which in turn indicates the role of cell or clone extrinsic processes in mediating the influence of EPO on CH. These findings support the hypothesis that EPO may selectively promote the competitive fitness of healthy HSPC clones over those with pathogenic mutations in *DNMT3A* and *TET2* that otherwise have a growth advantage. EPO exerts its effects through binding to the EPO receptor on HSPCs, activating downstream signalling pathways such as JAK2/STAT5, PI3K/AKT, and MAPK/ERK, which are crucial for cell survival, proliferation, and differentiation^28^. Additionally, EPO may influence the bone marrow microenvironment, modulating factors such as hypoxia or resistance to hypoxia and inflammation that can impact HSPC behaviour and clonal selection. Specifically, EPO has been found to inhibit the pro-inflammatory NF-kB pathway and downregulate the production of pro-inflammatory cytokines such as IL-6^29^.

Our findings have potential clinical implications since erythropoiesis stimulating agents (ESAs), which are synthetic recombinant forms of EPO, are widely available. If elevated EPO levels are indeed protective against CH, therapeutic strategies aimed at modulating EPO levels or enhancing EPO signalling could be explored to prevent or delay the onset of CH and its progression to haematological malignancies. Such interventions may provide maximal benefit in individuals with low EPO levels, or conditions which cause EPO resistance (e.g. iron and vitamin D deficiencies, inflammation, and secondary hyperparathyroidism) or impair EPO production (e.g. chronic kidney disease). Interestingly, chronic kidney disease progression and lower glomerular filtration rate have been associated with CH, which was supported by MR analyses amongst diabetic individuals^30^. Consistent with this observation we also found that the common variant used to proxy plasma EPO levels in our study was associated with diabetic nephropathy. As serum EPO levels decline steadily with renal dysfunction it is plausible that EPO deficiency may therefore contribute to development of CHIP in chronic kidney disease. However, caution with EPO modulation is warranted given the known risks associated with exogenous EPO administration, such as thrombosis and hypertension^31^. Any potential intervention would require rigorous clinical evaluation to balance efficacy with safety. Two multiparameter clinical risk prediction models, CHRS^32^ and MN-Predict^33^, have recently been introduced to predict the risk of progression to myeloid cancers in the setting of CH and there is scope, pending further clinical investigation, for interventions such as ESAs to be used to manage CH in those predicted to be at high risk for progression to cancer.

Our study has several strengths. By leveraging the random assortment of alleles at conception, the use of MR minimises the likelihood of confounding and reverse causation, which commonly afflict observational studies. We used rs1617640 as the primary genetic instrument in the *EPO* gene region, given that this promoter variant (i) has been shown to be associated with plasma EPO levels in two prior GWAS^13, 16^, (ii) is associated with EPO gene expression^16^, and (iii) editing rs1617640 in isogenic embryonic human kidney cells altered expression of EPO and of Notch signalling genes (*HEY2, DTX3L, PARP9*) known to be downstream targets of *EPO*^16^. Furthermore, we used the largest GWAS of plasma EPO levels and for CH risks in our MR analyses. Our Bayesian re-analysis offered a probabilistic interpretation that avoided dichotomisation of results at a P < 0.05 threshold and indicated strong support that elevated EPO levels are protective against CH under a range of priors. Finally, additional sensitivity analyses demonstrated that the association was not being driven by sample overlap between the exposure and outcome GWAS.

Our findings should be interpreted in the context of some limitations. First, our analyses were predominantly conducted in individuals of European ancestry, which may limit the generalisability of findings to other populations. While we observed consistent effect directions in African, East Asian, and South Asian ancestries, the estimates were imprecise due to smaller sample sizes. Second, the availability of only single variants that explain just a small fraction of the variance in plasma EPO levels as instruments for EPO may have attenuated the observed associations due to lack of power. Third, in “drug target” MR analyses such as ours which are based on *cis*-regulatory/promoter variants or exonic mutations, statistically ruling out pleiotropy and methodologically evaluating the exclusion-restriction assumption, which states that the genetic instruments affect the outcome only through the exposure, is challenging^34-36^. However, given our use of variants in the *EPO* gene as instruments, biologically it is highly likely that these directly influence plasma EPO protein levels. Moreover, our search in PhenoScanner did not reveal any significant pleiotropic associations that could confound our results. Fourth, while colocalisation analyses suggested that there was reduced potential for bias from linkage disequilibrium between distinct variants underlying the genetic associations for plasma EPO levels and CH risk at the EPO locus, the lack of power in these analyses have limited our ability to draw definitive conclusions. Fifth, plasma EPO concentrations may not fully capture the local EPO activity within the bone marrow, where HSPCs reside.

In conclusion, we report evidence that suggests a protective role for elevated germline genetically predicted plasma EPO levels against overall, *DNMT3A*- and *TET2*-mutant CH. These findings contribute to the understanding of the aetiology of clonal haematopoiesis with potential implications for progression to haematological malignancies and the broader landscape of ageing and disease. Tailoring EPO administration to individuals at high risk of developing CH and progression to malignancy may open fresh avenues for personalised myeloid cancer prevention. Further research studies, including clinical trials, are essential to validate these associations and explore the potential of targeting EPO in CH prevention and management.

## Data Availability

The data used in this study is publicly available on Zenodo at https://zenodo.org/records/6811853 and on the GWAS Catalog at https://www.ebi.ac.uk/gwas/downloads/summary-statistics.

https://www.ebi.ac.uk/gwas/downloads/summary-statistics

## CODE AVAILABILITY

The code used in this study can be found at: https://github.com/GeorgeRichenberg/EPO_CH_MR

## ETHICS DECLARATIONS/COMPETING INTERESTS

TRG receives funding from Biogen and GSK for unrelated research. The other authors declare no competing interests.

## CONTRIBUTIONS

G.R. and S.P.K. conceived and designed the study. G.R. performed the data analyses and visualisation. T.R.G. and P.C. were involved in data interpretation. G.R. wrote the first draft of the manuscript which was then edited by S.P.K. All authors critically reviewed the manuscript and contributed important intellectual content. All authors have read and approved the final manuscript as submitted.

## FIGURE LEGENDS

**Supplementary Figure 1:** MR results for the EPO and overall, *DNMT3A*-, and *TET2*-mutant CH risk associations using variant-EPO association estimates from a GWAS of 6,127 individuals that did not include the UK Biobank (labelled as “non-UKB” and compared to corresponding “UKB”-based associations).

