## Supplementary figures and images for "Genetically predicted plasma erythropoietin levels and clonal haematopoiesis risk: a Mendelian randomisation study"

### Supplementary Figure 1

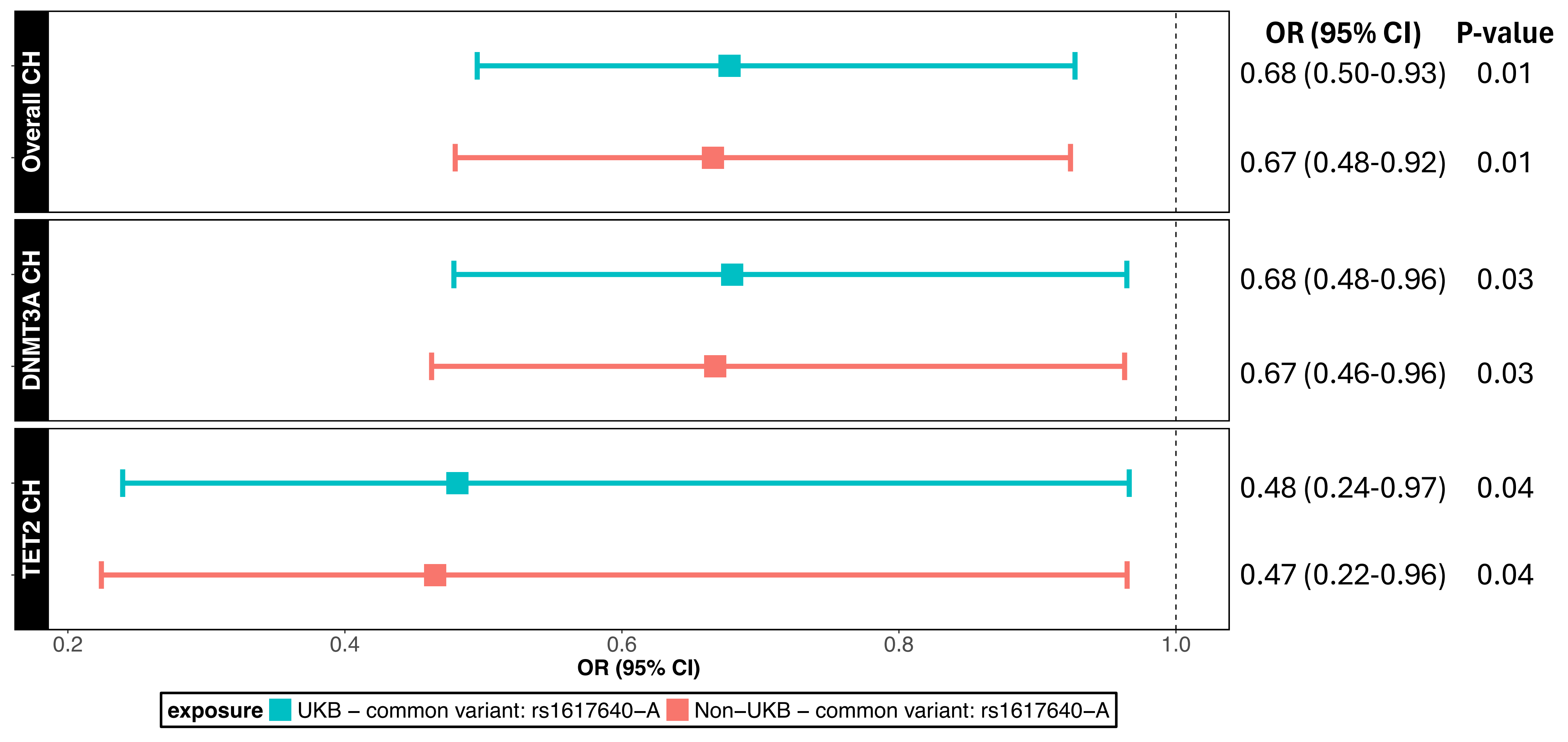
